# Population based mean Vitamin D levels in 19 European Countries & COVID-19 Mortality

**DOI:** 10.1101/2021.03.11.21253361

**Authors:** Amar Ahmad, Christian Heumann, Raghib Ali, Tim Oliver

## Abstract

**Objectives:** Reports early in the epidemic linking low mean national Vitamin D level with increased COVID-19 death, and until recently little research on the impact of Vitamin D deficiency on severity of COVID-19, led to this update of the initial report studying mortality up to the end of January 2021.

**Design and Setting:** Coronavirus pandemic data for 19 European countries were downloaded from Our World in Data, which was last updated on January 24, 2021. Data from March 21, 2020 to January 22, 2021 were included in the statistical analysis. Vitamin-D (25)-HD mean data were collected by literature review. Poisson mixed-effect model was used to model the data.

**Results:** European countries with Vitamin-D (25)-HD mean less than or equal to 50 have higher COVID-19 death rates as compared with European countries with Vitamin-D (25)-HD mean greater than 50, relative risk of 2.155 (95% CI: 1.068 – 4.347, p-value = 0.032). A statistically significant negative moderate Spearman rank correlation was observed between Vitamin-D (25)-HD mean and the number of COVID-19 deaths for each 14-day period during the COVID-19 pandemic time period.

**Conclusions:** The observation of the significantly lower COVID-19 mortality rates in countries with lowest annual sun exposure but highest mean Vitamin-D (25)-HD levels provides support for the use of food fortification. The need to consider re-configuring vaccine strategy due to emergence of large number of COVID-19 variants provides an opportunity to undertake such therapeutic randomized control trials.

## INTRODUCTION

The COVID-19 pandemic has created a major global health problem because of its high number of patients requiring intensive care and death rate particularly in patients over 65-70 yrs of age. (1-3) The impact has been unequal globally, primarily due to the younger populations in, for example, Africa where the majority of the population are under 65 (4) not getting ill, except for South Africa (5). Non-pharmaceutical interventions used to control transmission, particularly universal lockdowns have been the primary means of reducing case incidence and this has caused serious economic harm in many countries (6-8).

Vitamin-D has a potential role in infections, which is implied from its effect on the innate and adaptive immune responses (9) and the fact that respiratory infections tend to disappear during summer. Furthermore, there is existing evidence for an association between vitamin-D deficiency and a risk of influenza infection and when the individuals have Vitamin D deficiency, they have reduced infection when entered into supplementation randomized trials (10, 11), though dose and scheduling is far from clear.

In the 12 months since the onset of the COVID-19 pandemic, no major trials of Vitamin D have been completed. There has however been increasing interest with 5 reports demonstrating a strong impact of Vitamin D deficiency on the behavior of COVID-19 illness (12-16) and limited data from three small randomized trial reporting improved clinical outcome in patients receiving replacement therapy (17) that has been followed by posting on-line of a 930 cluster randomized trial with Calciferol reporting a 64% reducation in deaths.

Population-based mean levels of vitamin-D were shown in an analysis duting the first 2 months of the pandemic to correlate inversely with COVID-19 mortality (18). The aim of this paper is to update these analyses after 12 months of the pandemic through statistical analysis focusing on the association between vitamin D (25)-HD average and COVID-19 mortality rates based on public record of the number of COVID-19 deaths in 2-week periods from 21/02/2020 – 22/01/2021 in 19 European countries.

## STUDY DESIGN

Coronavirus pandemic data for 19 European countries were downloaded from “Our World in Data” (see https://ourworldindata.org/coronavirus), which was last updated on January 24, 2021 (19). Data from March 21, 2020 to January 22, 2021 were included in the statistical analysis. Data on vitamin D deficiency were collected via a literature review (18, 20, 21).

## STATISTICAL ANALYSIS

Last observation carried forward was used to impute missing values, whenever data were not available (22). The vitamin D mean value (18) was missing for Greece which was (single) imputed by the median vitamin D value of all countries.

The crude mortality rate (CMR) was computed using data from January 22, 2021 by dividing the total number of COVID-19 deaths by the corresponding population per 100,000. Spearman rank correlation was estimated between vitamin D averages variable and the number of COVID-19 total deaths and the crude mortality rate respectively. A jackknife empirical 95% confidence interval for Spearman’s correlation was computed (23).

Then the vitamin D values were binarised into 0 (vitamin D ≤ 50) versus 1 (vitamin D > 50). Wilcoxon rank-sum test was used to compare the total number of COVID-19 deaths and CMR between countries who have vitamin D averages of less than or equals to 50 versus countries that have vitamin D averages greater than 50 respectively.

Univariate and bivariate generalised linear regression models with Quasi-Poisson distribution were performed using the total number of COVID-19 deaths by January 22^nd^ 2021 as an outcome. The predictors were the categorical vitamin D variable and the proportion of age 70+ for each country. The population variable was added as an offset in the model. Relative risks (RRs) and 95% confidence intervals (95% CI) were computed with corresponding z- and p-values.

A time-point variable of 22 time-periods of 14 days was created between 01/03/2020 and 22/01/2021 at each time-point. A Spearman rank correlation was used to evaluate the association between the number of COVID-19 deaths at each time-point and the average of vitamin D of the 19 European countries. To compute an approximate 95% confidence interval (using the normality approximation) for the estimated Spearman rank correlation, a leave-one-time-period-out cross-validation was performed at each time-point.

A Poisson mixed effects regression model (24) was fitted with the total number of new COVID-19 deaths as an outcome. The percentage of age 70+ and binarized vitamin D were used as fixed effect predictors. The time-periods and the 19 European countries variable were used as random effect variables with (04/04/2020 to 17/04/2020) and the UK as reference groups respectively. The populations variable was added as an offset in the model. Relative risks (RRs) and 95% confidence intervals (95% CI) were computed with corresponding z- and p-values.

All applied statistical tests were two-sided, p-value < 0.05 were considered statistically significant. Statistical analyses were performed in R version 4.0.2 (25)(25)(25).

## RESULTS

The data used in this statistical analysis are shown in Table 1 in descending order of the countries level of Vitamin D level. It can be seen visually that the crude mortality rate (CMR) increases with decreasing mean Vitamin D level. A statistically significant moderate negative Spearman’s ρ correlation was observed between the total number of COVID-19 deaths and the average of vitamin D, ρ = -0.516 (95% CI: -0.860 – -0.168) as well as between COVID-19 crude mortality rates and the average of vitamin D, ρ = -0.430 (95% CI: -0.805 – -0.081) (Figure 1). A similar result was seen in the numbers of Table 2. This shows the Spearman’s rho correlation analysis between the number of new COVID-19 deaths at each of the 22 time periods and the same reported mean vitamin D used on each occasion. All the observed negative Spearman’s rho correlation values were statistically significant.

**Table 1:** Data used in the statistical analysis. Data sorted by vitamin-D values

|  |  |  |  |  |  |
| --- | --- | --- | --- | --- | --- |
| Country | Total Deaths | Population | Age 70+ | Vitamin D | CMR |
| Portugal | 9920 | 10196707 | 14.924 | 39.0 | 97.3 |
| Spain | 55441 | 46754783 | 13.799 | 42.5 | 118.6 |
| Switzerland | 9034 | 8654618 | 12.644 | 46.0 | 104.4 |
| UK | 96166 | 67886004 | 12.527 | 47.4 | 141.7 |
| Belgium | 20675 | 11589616 | 12.849 | 49.3 | 178.4 |
| Italy | 84674 | 60461828 | 16.24 | 50.0 | 140 |
| Germany | 51713 | 83783945 | 15.957 | 50.1 | 61.7 |
| Austria | 7330 | 9006400 | 13.748 | 56.0 | 81.4 |
| Ireland | 2870 | 4937796 | 8.678 | 56.4 | 58.1 |
| Greece | 5598 | 10423056 | 14.524 | 57.95 | 53.7 |
| Netherlands | 13528 | 17134873 | 11.881 | 59.5 | 79.0 |
| France | 72788 | 65273512 | 13.079 | 60.0 | 111.5 |
| Hungary | 11811 | 9660350 | 11.976 | 60.6 | 122.3 |
| Czechia | 15130 | 10708982 | 11.58 | 62.5 | 141.3 |
| Denmark | 1942 | 5792203 | 12.325 | 65.0 | 33.5 |
| Norway | 544 | 5421242 | 10.813 | 65.0 | 10.0 |
| Finland | 644 | 5540718 | 13.264 | 67.7 | 11.6 |
| Sweden | 11005 | 10099270 | 13.433 | 73.5 | 109.0 |
| Slovakia | 3894 | 5459643 | 9.167 | 81.5 | 71.3 |

**Table 2:** Spearman correlation (95% CI) between vitamin-D and the new number of COVID-19 deaths at different time-points

|  |  |  |
| --- | --- | --- |
| Time-Point | Date | Spearman $\rho$ (95% CI) |
| 1 | 21/03/2020 – 03/04/2020 | -0.571 (-0.585, -0.553) |
| 2 | 04/04/2020 – 17/04/2020 | -0.614 (-0.629, -0.595) |
| 3 | 18/04/2020 – 01/05/2020 | -0.543 (-0.559, -0.525) |
| 4 | 02/05/2020 – 15/05/2020 | -0.478 (-0.496, -0.457) |
| 5 | 16/05/2020 – 29/05/2020 | -0.457 (-0.476, -0.436) |
| 6 | 30/05/2020 – 12/06/2020 | -0.285 (-0.309, -0.261) |
| 7 | 13/06/2020 – 26/06/2020 | -0.341 (-0.360, -0.320) |
| 8 | 27/06/2020 – 10/07/2020 | -0.431 (-0.448, -0.412) |
| 9 | 11/07/2020 – 24/07/2020 | -0.421 (-0.437, -0.403) |
| 10 | 25/07/2020 – 07/08/2020 | -0.365 (-0.378, -0.349) |
| 11 | 08/08/2020 – 21/08/2020 | -0.464 (-0.477, -0.449) |
| 12 | 22/08/2020 – 04/09/2020 | -0.443 (-0.455, -0.428) |
| 13 | 05/09/2020 – 18/09/2020 | -0.465 (-0.480, -0.447) |
| 14 | 19/09/2020 – 02/10/2020 | -0.395 (-0.413, -0.375) |
| 15 | 03/10/2020 – 16/10/2020 | -0.461 (-0.480, -0.439) |
| 16 | 17/10/2020 – 30/10/2020 | -0.457 (-0.475, -0.437) |
| 17 | 31/10/2020 – 13/11/2020 | -0.489 (-0.505, -0.471) |
| 18 | 14/11/2020 – 27/11/2020 | -0.508 (-0.523, -0.490) |
| 19 | 28/11/2020 – 11/12/2020 | -0.457 (-0.471, -0.440) |
| 20 | 13/12/2020 – 25/11/2020 | -0.447 (-0.462, -0.429) |
| 21 | 26/12/2020 – 08/01/2021 | -0.385 (-0.401, -0.366) |
| 22 | 09/01/2021 – 22/01/2021 | -0.405 (-0.421, -0.387) |

**Figure 1:**
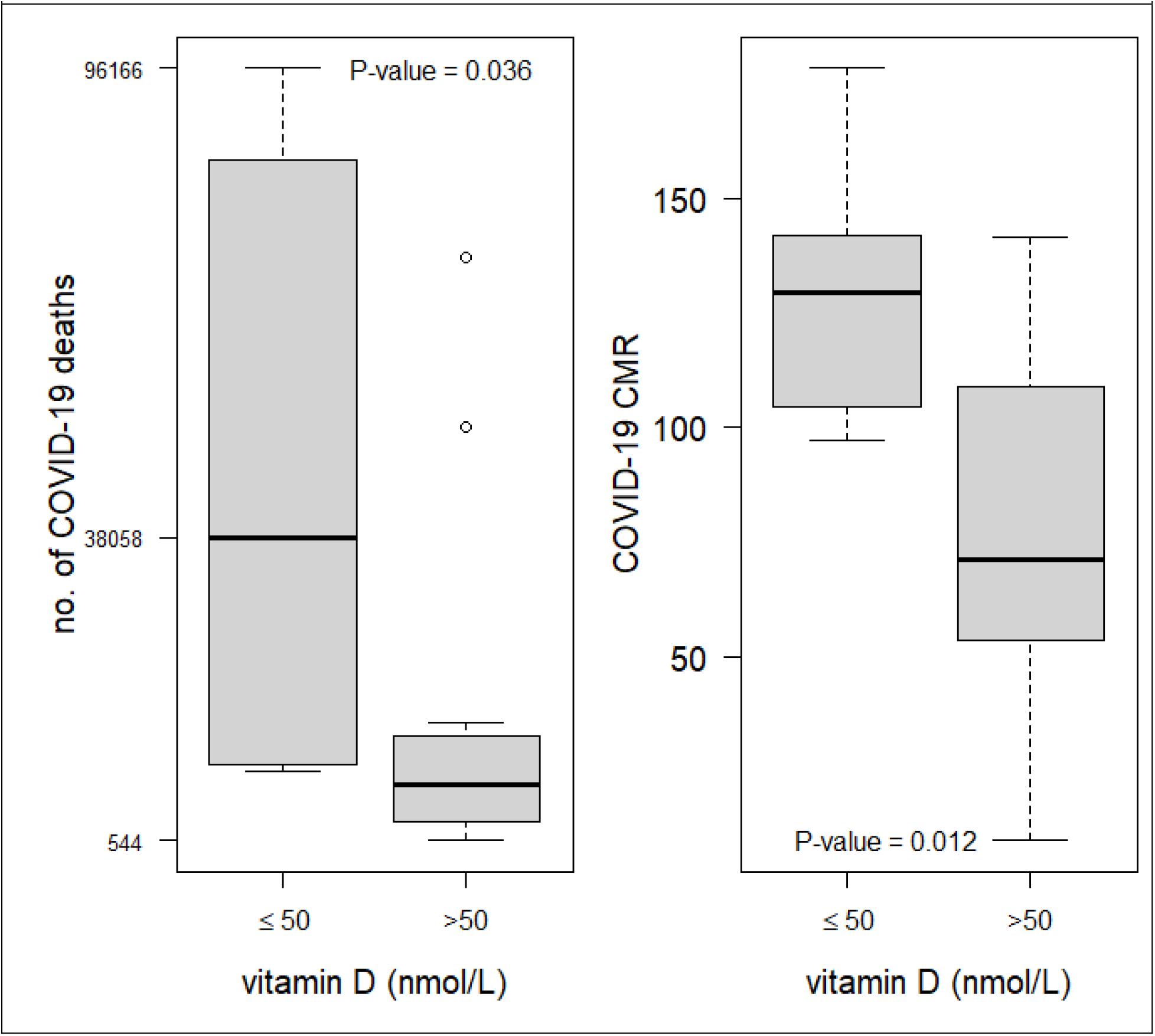
Distribution of the total number of COVID-19 deaths with the vitamin-D group.

Figure 1 shows the distribution of the total number of COVID-19 deaths and CMR within the vitamin D group (≤50 versus >50) respectively. A statistically significant difference was observed between the vitamin D groups (≤50 versus >50), Wilcoxon sum-rank test p-value of 0.036 and 0.012 respectively.

Table 3 shows the results of the univariate and bivariate generalised linear regression model with family quasi-Poisson using the total number of COVID-19 deaths from 22/03/2020 as an outcome. The countries with vitamin D average ≤ 50 have higher COVID-19 death rates as compared with countries with vitamin D average > 50, RR of 1.642 (95% CI 1.274 – 2.118, p-value = 0.02) univariately. This result holds after adjusting for the population age structure (ie percentage of the population with age 70+), RR of 1.663 (95% CI: 1.293 – 2.140, p-value < 0.0001). This result is similar to the result from the fitted Poisson mixed-effects models (Table 4), where countries with vitamin D average ≤ 50 have higher COVID-19 death rates as compared with countries with vitamin D average > 50 before and after adjusting for population age structure with RR of 2.197 (95% CI: 1.131 – 4.271, p-value = 0.02) and 2.155 (95% CI: 1.068 – 4.347, p-value = 0.032) respectively. The percentage of age 70+ was not statistically significant in either a univariate and bivariate regression models (Table 3 and 4).

**Table 3:** Univariate and bivariate quasi-Poisson regression models. The percentage of age 70+ and binarized vitamin D variables were added together

|  | Univariate |  | Bivariate |  |
| --- | --- | --- | --- | --- |
| Variable | RR (95% CI) | z-value (P) | RR (95% CI) | z-value (P) |
| age 70+ | 0.978 (0.892, 1.073) | -0.471 (0.637) | 0.964 (0.896, 1.036) | -1.001 (0.317) |
| vitamin D ≥ 50 | Reference |  |  |  |
| vitamin D < 50 | 1.642 (1.274, 2.118) | 3.823 (0.02) | 1.663 (1.293, 2.140) | 3.957 (<0.001) |

**Table 4:** Poisson mixed effect regression models. Univariate models, where fitted with single fixed effect variable. The percentage of age 70+ and binarized vitamin D variables were added together as fix-effect predictors.

|  | Univariate | Bivariate |
| --- | --- | --- |

| Variable | RR (95% CI) | z-value (P) | RR (95% CI) | z-value (P) |
| --- | --- | --- | --- | --- |
| age 70+ | 1.084 (0.921, 1.276) | 0.972 (0.331) | 1.013 (0.867, 1.183) | 0.165 (0.869) |
| vitamin D ≥ 50 | Reference | | | |
| vitamin D < 50 | 2.197 (1.131, 4.271) | 2.322 (0.020) | 2.155 (1.068, 4.347) | 2.145 (0.032) |

## CONCLUSION

Despite regularly assuring the UK population that the pandemic would lessen during our summer months, there has been no funding for clinical trials of Vitamin D supplements and negative reviews mainly based on others data by PHE, SACN, NICE (https://www.nice.org.uk/guidance/ng187). This negative view focuses attention onto an on-going controversy dating back to the 1950s, i.e. whether food should be supplemented with Vitamin D. In the early years after the 2^nd^ world war in the UK enthusiasm for supplementation was so great that children died from overdose hypercalcaemia. As a result, the UK has been more reluctant to sanction such supplementation while it has been used increasingly used in the Scandinavian countries (26). This could explain why most of these countries, despite having less sunshine than the UK, have lower RRs. The increasing data to support this interpretation started with Meltzer et al studying Vitamin D levels in 489 attendees in an urban medical centre in Chicago (13). They demonstrated a higher frequency of acquisition of COVID-19 cases in the Vitamin D deficient group (21.9%) than in the sufficient group (12.2%). This was subsequently confirmed by a much larger study (14) and two smaller studies (12, 15). The Boston group also reported data suggesting a link between Vitamin D deficiency and increased severity of disease (16).

In a recent systematic review and meta-analysis, Pereira et al (27) show, that vitamin D insufficiency increased COVID-19 mortality (OR = 1.82, 95% CI = 1.06–2.58).

Using UK Biobank data of 502,624 participants aged 37–73 years between 2006 and 2010, Hastie et al (28, 29) show that COVID-19 participants who had COVID-19 have lower vitamin-D level with median of 43.8 (IQR: 28.7-61.6) as compared with those who had no COVID-19 median of 47.2 (IQR: 32.7–62.7), Wilcoxon’s p-value < 0.01. However, this result was not statistically significant after adjustment for confounders, though this did not take allowance for the fact that some of the confounders were also associated with Vitamin D deficiency.

In a cohort of 185 patients at the Medical University Hospital Heidelberg-Germany, vitamin-D deficiency was associated with higher risk of invasive mechanical ventilation and deaths after adjusting for age, gender, and comorbidities, HR of 6.12 (95% CI: 2.79–13.42, p < 0.001) and 14.73 (95% CI 4.16–52.19, p < 0.001) (30, 31).

Karahan and Katkat (32) show that vitamin-D insufficiency was present in 93.1% of the patients with severe-critical COVID-19, and that vitamin-D 25(OH) mean was significantly lower in patients with severe-critical COVID-19 compared with moderate COVID-19.

Despite all of these strong indications that Vitamin D could play an important role in the control of the COVID-19 pandemic, formally published papers have less than 500 patients recruited to randomized trials of Vitamin D(27). There has been one prospective observational study on 410 patients from India which at first sight was considered as a failure of Vitamin D supplementation (https://www.researchsquare.com/article/rs-129238/v1, currently undergoing major revision). As it gave Vitamin D to two thirds of those with Vitamin D deficiency who were also younger than those who weren’t, the inverse of what is expected from other studies and overall there was a very low mortality rate, there was no difference in survival between those with and those without Vitamin D deficiency.

No data has been published from the UK about the effect of Vitamin D replacement. This is despite a continuous debate since April 2020 about Vitamin D deficiency contributing to excess deaths in the BAME community (https://www.gov.uk/government/publications/covid-19-understanding-the-impact-on-bame-communities). Throughout the year, the BAME community are known to have 20% less circulating vitamin D (33) and even in summer more than 30% have severe deficiency compared to less than 6% in white European population (34) and probably have done so for many of the years since birth. In addition, there are two studies, one a randomized trial, that have clearly demonstrated that they need 10 times more Vitamin D than recommended in the SACN report (35, 36).

Clearly, accelerating the role out of Vaccines around the world should not diminish. However as our data could not adjust for well established COVID-19 confounders such as Diabetes, Obesity, social deprivation and poverty, the increasing availability of simple finger-prick techniques (www.vitamindtest.org.uk) that speed up measuring of Vitamin D should make better selection for such therapy trials easier to recruit. Given reports published on line but still under review (see https://www.imperial.ac.uk/media/imperial-college/institute-of-global-health-innovation/REACT-2-round-5-preprint.pdf table 4) that the over 70’s population have a 62% failure rate to produce IgG antibodies >21 days after first vaccination compared to 24% in 50-69 year olds and 7% in <50 year olds and the older age groups have a well recognized risk of lower Vitamin D levels, there could be a strong case to evaluate Vitamin D supplements in such patients.

## Conclusion

The data from this statistical analysis shows a strong and statistically significant association between the Vitamin D deficiency and the total number of COVID-19 deaths in the 19 European countries included in this statistical analysis. The new vitamin D techniques for easier detection of deficiency using the finger prick technology should enable better selection of patients to benefit from treatment and prove it with .appropriately selected patients in treatment trials and reduce poor immune response in vaccine recipients

## Data Availability

Data available through Amar Ahmad

